# Trends in Technology Usage for Parkinson’s Disease Assessment: A Systematic Review

**DOI:** 10.1101/2021.02.01.21250939

**Authors:** Ranadeep Deb, Ganapati Bhat, Sizhe An, Holly Shill, Umit Y. Ogras

## Abstract

Parkinson’s disease (PD) is a neurological disorder with complicated and disabling motor and non-motor symptoms. The complexity of PD pathology is amplified further due to its dependency on patient diaries and the neurologist’s subjective assessment of clinical scales. This challenge can be addressed by the advances in mobile technology, which can enable objective, accurate, and continuous patient monitoring. Indeed, a significant amount of recent work explores new cost-effective and subjective assessment methods of PD symptoms. For example, smart technologies, such as wearable sensors, have been used to analyze a PD patients’ symptoms to assess their disease progression and even to detect signs in their nascent stage for early diagnosis of PD.

This review focuses on the use of modern wearable and mobile equipment for PD applications in the last decade. Four significant fields of research were identified: Assistance to Diagnosis, Prognosis or Monitoring of Symptoms and their Severity, Predicting Response to Treatment, and Assistance to Therapy or Rehabilitation. This study starts with 31,940 articles published between January 2008 and December 2019 in the following four databases: Pubmed Central, Science Direct, IEEE Xplore and MDPI. A total of 976 papers are manually investigated and included in this review after removing unrelated articles, duplicate entries, publications in languages other than English, and other articles that did not fulfill the selection criteria. Our analysis shows that the numbers of published papers every year has increased at a constant rate from 2008 to 2015, while the rate of increase has significantly grown from 2016 to 2019. Majority of the papers (62%) were published in the last four years, and 21% papers in just 2019. In terms of the symptoms, gait and tremor are two major ones that researchers have focused on. The trend shows the growing interest in assessing Parkinson’s Disease with wearable devices in the last decade, particularly in the last 4 years. Our automated script makes the review easily reproducible for publications published in the future.

## I. Introduction

Parkinson’s disease (PD) is a complex neurodegenerative disorder that affects patients’ and their caregivers’ overall quality of life (QoL). Approximately 60,000 individuals in the United States are diagnosed with PD each year, while more than 10 million people are living with PD worldwide [1], [2]. PD is often seen together with many significant motor signs, such as tremor, rigidity, bradykinesia, hypokinesia, postural instability, and gait difficulties. While its clinical diagnosis is usually based on these motor symptoms, many non-motor symptoms also manifest themselves with the disease. These non-motor symptoms are commonly evident and sometimes more disabling than motor symptoms. Common non-motor signs of the disease are cognitive impairment, reduced ability to smell, dementia, depression, and emotional changes. It is a progressive disorder whose symptoms become more noticeable with age.

The current practice for assessing the motor and non-motor symptoms of PD patients is a neurological examination, during which a neurologist watches the patient perform specific tasks. Neurologists assign scores to the tasks performed by the patient as described by the Unified Parkinson’s Disease Rating Scale (UPDRS) [3] or its updated version, the Movement Disorder Society-sponsored revision of the UPDRS (MDS-UPDRS) [4]. Another rating scale, the Hoehn and Yahr scale (HY) [5] assigns an overall score out of 5 to the patient based on their clinical stage. These clinical scales are subjective and can lead to high inter-rater variability among neurologists and clinics. Similarly, the clinical assessment relies also on the symptoms’ progression described in the patient’s diary. The credibility of such reports is limited by subjectivity and recall bias of the patient [6], [7]. Second, the cost of current treatment approaches is high. Medication alone can cost $2,500 a year, while a corrective surgery, like Deep Brain Stimulation (DBS), costs up to $100,000 per person [1]. For example, imaging equipment, such as magnetic resonance imaging (MRI), single-photon emission computed tomography (SPECT), and positron emission tomography (PET), are used to assist the neurologist in making an objective and more accurate diagnosis [8]. High equipment costs factor into the expenses of diagnosis and treatment of Parkinson’s Disease [9].

Advances in mobile computing technologies facilitate long-term objective measurement of symptoms. Hence, new systems based on mobile technology can enable a wide range of monitoring, diagnosis, and rehabilitation applications [10]–[14]. Indeed, the number of publications that report using wearable and mobile technology for PD research increased by 100 times between 2008 and 2019, as the results presented in Section V. Popular devices include inertial measurement units (IMUs), force and pressure plates, biopotential sensors, and optical motion capturing systems. IMUs usually include accelerometer and gyroscope sensors, which can record essential data for analyzing the symptoms. Similarly, force sensors in a force plate provide information about the patient’s posture and balance. As a complementary modality, electroencephalogram (EEG) and electromyogram (EMG) sensors measure neural activity and muscular response, respectively. In contrast, optical motion capturing systems like VICON and Microsoft Kinect analyze patients’ body motion in their ambulatory environment. Interconnection of these sensors has also become straightforward with the growing use of communication protocols, such as Zigbee and Bluetooth.

A large number of recent studies investigated the use of wearable sensors and other technologies to assess the symptoms of a patient suffering from neurological disorders [15]. There is also a growing interest in getting an unbiased analysis of the efficacy of technology-based devices that can be used in scientific research of health monitoring and clinical practices [16]. For example, [16] reviewed 168 articles after searching the PubMed database and grouped the studies based on the type of device used. They classified the devices as (i) ‘recommended’, (ii) ‘suggested’ or (iii) ‘listed’ based on the following criteria: (1) used in the assessment of Parkinson’s disease, (2) used in published studies by people other than the developers, and (3) successful clinimetric testing. They concluded that objective sensing technology is gaining attention in the study of Parkinson’s Disease, but the clinimetric properties and testing of the devices remain a controversy. They surmised that PD symptoms like postural control, bradykinesia, tremor, freezing, dyskinesia, gait, and daily activity/physical activity could be objectively measured using the reviewed devices. Applications of wearable technology to the assessment of PD symptoms has also increased in recent years [17]. Wearable sensors have shown promise in PD diagnosis and management, as well as for other pathology [18]–[21]. For example, [21] discuss how wearable and mobile technologies could improve the management of Essential Tremor (ET), one of the most common movement disorders that impact millions of people worldwide. The study proposed 7 different areas in which mobile and wearable technology can improve the clinical management of ET and review the current state of research in these areas. The authors conclude that the presence of mobile and wearable technology everywhere could be leveraged to improve the quality of life of the patients if clinicians, engineers, and computer scientists work together on addressing the current knowledge gaps.

This paper presents a systematic review of mobile technology use in the PD context by analyzing articles published between January 1, 2008, and December 31, 2019. An electronic database search in Pubmed Central, Science Direct, IEEE Xplore, MDPI is performed to retrieve all articles related to Parkinson’s Disease (with the keyword “Parkinson”). This search yields 25,600 articles after removing surveys and articles written in a language other than English. Then, the articles are filtered by applying one blocks of keywords related to the disease, such as “Tremor” and “Freezing of gait (FoG)”. Similarly, the remaining blocks of keywords filter articles that employ non-human subjects, out of scope technologies, and devices, as detailed in Section IV-B. After the automatic filtering using the keywords, these articles are inspected and classified manually to identify the following aspects: (1) the application areas of the proposed solution, (2) the symptoms measured for the application, (3) the devices used for measuring the symptoms, and (4) the sensors included in the devices. The papers under study and our classification are saved in a spreadsheet and released to the public at https://bit.ly/2HVwKZ0. The spreadsheet can be updated as new solutions are developed in the future. Hence, it can serve as a platform for technology professionals and clinicians to find innovative developments in the area. Furthermore, this spreadsheet can be used by researchers working in this area to compare their work with the other existing work in their field. The manually labeled data from this spreadsheet can also enable automatic classification of papers studying PD symptoms with modern devices.

We have also developed an automated filtering framework for articles focusing on PD assessment with novel technologies based on their content in Title, Abstract, and Keywords (TAK). The filtering criteria are developed following the Preferred Reporting Items for Systematic Reviews and Meta-Analyses (PRISMA) standard [22]. Our criteria also follow the PICO (Patient, Intervention, Comparison, Outcome) principles [23]. Manual filtering is limited by time and subjectivity, which can be avoided by our filtering framework. It can objectively select the relevant articles for any systematic review or find the related literature in a specific field of work. It can also detect duplicate items across multiple databases and filter them out.

*The automated script makes the review easily reproducible for publications published in the future*.

We then reviewed each article as follows:

- Analyze the trends in the use of technology for PD research. For example, we use the data to identify the most popular application areas and the areas that receive increasing attention (Section II),
- Analyze the devices used in each application area, and how the use of devices has progressed in the last decade (Section III),
- Analyze the symptoms measured in each application area and determine the most commonly used devices for measuring the different traits (Section V-A),
- Examine the overall trend of research in this field and how it progresses in the coming years (Section V-B, Section V-C).

## II. Application Areas

Mobile technology can be used to track symptoms that can be used to assess the progression of a PD patient. These symptoms, referred to as cardinal PD features, are divided into motor symptoms and non-motor symptoms, as shown in Table I. The motor symptoms are the movement impairments caused due to Parkinson’s Disease. The motor symptoms are traditionally considered as tremor, rigidity, bradykinesia and postural instability. Additionally, abnormalities in limb movements like hand rotation, finger tapping, and arm angle etc., are also considered as motor symptoms. In contrast, non-motor symptoms include a large variety of cardinal features, such as sleep disturbance, cognitive activity, fatigue, dementia, and psychiatric impairments, as listed in the second column of Table I. Finally, mixed symptoms, such as speech and swallowing, exhibit a combination of motor and non-motor activities. Figure 1 shows the number of papers that measure the different motor and non-motor symptoms. Gait abnormalities, including FoG are the most common symptoms measured, followed by tremor. Movement problems like bradykinesia and dyskinesia are also common symptoms in PD patients, and many research articles use modern measurement techniques to monitor these symptoms.

**TABLE I:** Different domains of assessment of Parkinson’s Disease using new technologies

| Signs and Symptoms |  |  |
| --- | --- | --- |
| Motor Signs and Symptoms | Non-motor Signs and Symptoms | Mixed Signs and Symptoms |
| Gait | EEG abnormalities |  |
| Limb movements | Cognitive activity |  |
| EMG abnormalities | Depression |  |
| FoG | Dementia |  |
| Tremor | Heart rate | Speech topics |
| Activities of Daily Living (ADL) | Emotions | Swallowing |
| Bradykinesia and Dyskinesia | Fatigue |  |
| Posture | Sleep topics |  |
| Balance | Blinking |  |
| Nocturnal Hypokinesia | Facial expression |  |
| Handwriting | Breath |  |
| Saccades | Cortical activity |  |

**Fig. 1:**
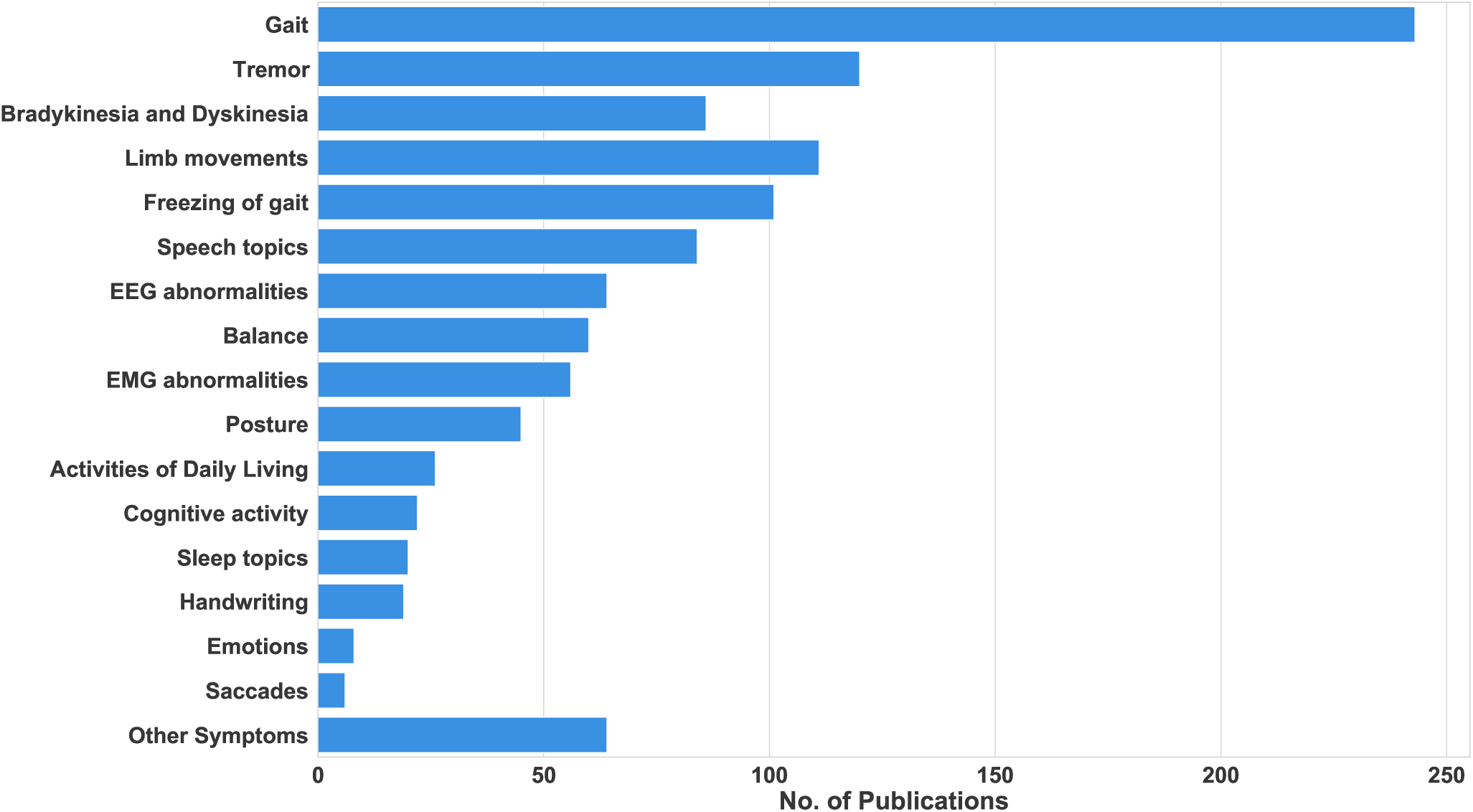
Number of publications between 2008-2019 that measure each symptom.

Mobile monitoring of motor and non-motor symptoms can be used in four major application areas in the PD research context: Diagnosis, Prognosis/Monitoring the Severity of Symptoms, Predicting the Response to Treatment, and Rehabilitation. These application areas are chosen based on previous reviews on PD [8], [17]. The rest of this section reviews these application areas.

### A. Diagnosis

The diagnosis of Parkinson’s Disease relies on the clinical assessment of the motor and non-motor symptoms by neurologists [24]. Neurologists observe the patients while they perform specific tasks. Then, they assign a score according to one of the standard scales: Unified Parkinson’s Disease Rating Scale (UPDRS) [3], its updated version, the Movement Disorder Society-sponsored revision of the UPDRS (MDS-UPDRS) [4], or the Hoehn & Yahr scale [5]. The clinical information derived from these rating scales is subjective. Hence, it leads to inter-rate variability and also intra-rate variability [8], [17]. Moreover, the equipment used to supplement the clinical assessment is expensive imaging tools like SPECT, PET, or MRI. Since early and accurate diagnosis is important, current techniques can be augmented with objective and cost-effective alternatives enabled by mobile technology. Therefore, recent work has used wearable sensors and other portable technology in the diagnosis of PD. These devices can provide objective measures for PD diagnosis that help in standardizing assessment [25]–[27]. Many researchers have also been able to use wearable sensors and other devices to differentiate PD patients and healthy controls in lab experiments [25], [26], [28].

In summary, mobile technology is used in the following sub-categories of PD diagnosis:

- Early diagnosis of patients with Parkinson’s disease
- Detecting Parkinsonian symptoms in people with untreated PD
- Differentiate patients with Parkinson’s disease from healthy controls or patients with a different neurological disorder
- Differentiate PD-related symptoms from similar symptoms but not caused by PD. For example, differentiating PD tremors from Essential Tremors (ET)

### B. Prognosis/Monitoring the Severity of Symptoms

Assessing the patient’s condition and severity of symptoms depend primarily on the clinicians’ judgment and the patient feedback from diaries and memory. The clinicians’ judgment is subjective [8], [17] while the patient’s diary and memory are limited by compliance and recall bias [6], [29], [30]. Since this approach may not be completely reliable, objective remote monitoring of PD symptoms is needed to assess disease progression, evaluate the severity of the symptoms, and continuously monitor the PD patients in unsupervised environments. To address these issues, recent work on PD prognosis focuses on the following areas:

- Home-based or remote monitoring of patients with PD
- Evaluating the progression of PD for a diagnosed patient
- Evaluating the severity of PD symptoms for a diagnosed patient

### C. Predicting Response to Treatment

To measure the efficacy of the treatment or the impact of the medication, clinicians rely on the patients’ recollection and their diaries, which can be subjective and unreliable. This problem motivated research on measuring the impact of a treatment, medication’s effectiveness in suppressing the PD symptoms, and assess their side effects. To this end, research on predicting response to treatment addresses the following issues:

- Measure the effect of treatment like Deep Brain Stimulation in suppressing the patient’s symptoms over time
- Measure the impact of a medication on the symptoms of the patient
- Measure the side effects of the medicine (e.g., levodopa induces dyskinesia)

### D. Rehabilitation

Physiotherapy and other rehabilitation techniques are among the most common treatments for movement disorders like Parkinson’s Disease. Like medication, it is crucial to assess their efficacy. Additionally, it has been observed that cues and feedback are beneficial in assisting a PD patient. A system can provide rhythmic auditory cues, visual cues, or haptic cues to facilitate a patient’s movement. Such a system can be used for gait training or to assess limb movements. Vibration-based actuators and audio feedback are useful to sensitize a patient suffering from Rigidity, Freezing of gait (FoG), Tremor, or other symptoms. They can also help patients break out of the freeze even suppress the symptom. Mobile technologies that target therapy and rehabilitation can be divided into the following sub-categories:

- Audio, visual, or haptic cue for gait or movement training
- Sensory feedback to suppress a symptom like FoG or Tremor

## III. Technology in Parkinson’s Disease Research

A number of device type and technologies are being used in the application areas presented in the previous section. We classify these devices into the following eight categories based on their form factor and sensing modalities.

### Wearable

Many recent approaches employ wearable devices for health monitoring since they facilitate recording patients’ activity and symptoms. Wearable devices are ideal for monitoring since they are not limited to a specific location and they can be easily integrated into patient’s clothes [31]. Most commonly used wearable devices contain sensors, such as inertial measurement units (IMUs). The outputs of these sensors can be processed to analyze the body movement, gait, and symptoms such as tremors. Our study includes the following wearable technologies:

1. IMUs with integrated accelerometer, gyroscope, and magnetometer sensors,
2. Insole sensors containing force or pressure sensors that can measure the Ground Reaction Force (GRF),
3. Wearable devices that can measure neural responses and muscle activities using sensors such as EEG and EMG,
4. Sensors incorporated with clothes or gloves, such as strain or accelerometer sensors to measure hand tremor,
5. Other wearable devices like smart glasses or smart hats which can record specific parameters like patient’s emotions.

### Biopotential Devices

Devices that can measure the electrical signal (biopotentials) that are generated by the physiological processes in our body. The tools that fall under this category are Electroencephalogram (EEG), Electrocardiogram, Electro-cardiogram, Magnetoencephalogram, and Electrooculogram. The biopotential devices can either be standalone, such as a a 16-lead EEG system, or in a wearable device with a single lead EMG sensor. When a biopotential device is integrated into a wearable device, it is included in both wearable and biopotential device categories.

### Cueing Devices

Devices that are used to give feedback or cues to the patients to rectify their walk or to assist with their movement fall under this category. Headphones or speakers can be used to deliver auditory cues, and visual cues can be provided using a screen or a smart glass or even in a virtual reality environment. Vibration sensors or electrical stimulators have been utilized to give sensory feedback to patients.

### Optical Motion Tracker

Devices that track motion of users with either radio frequency or optical signals fall under this category. For example, motion capturing systems, such as Microsoft Kinect and Vicon 3D, use structured light for analysis. Similarly, radar-based systems use radio frequency signals to monitor motion of subjects. A system of multiple IMUs, even camera-based 3D setups have also been used to capture the movement of a patient.

### Audio Recording

A number of PD patients experience issues in swallowing or in their speech, as shown in Table I. Audio recording devices are used to analyze and monitor symptoms related to speech and swallowing. For instance, speech recording devices, such as microphones and smartphones, are used to record the speech tasks or the patients’ voice to analyze the speech of the patient for diagnostic purposes [28], [32].

### Video Recording

Video recording devices provide utility for PD monitoring since gait and motor issues are one of the most common symptoms in PD. For instance, patients’ movements at home or during laboratory experiments are often recorded using a video camera to spot symptoms or to corroborate with predictions from other devices [33]–[35].

### Force/Pressure

Force or pressure plates measure the force exerted by the patients’ feet when they are walking. Therefore, they are useful to measure the quality of gait of the patient. Force or pressure sensors are also integrated into gait mats to measure the gait quality of a patient.

### Smartphone

More researchers are trying to use the sensors present on-board a smartphone (accelerometer, gyroscope, magnetometer, GPS) for different analyses. Smartphone applications are also used to record the patients’ mood, emotions and even to record the dosage or medicines. Additionally, the screen of the smartphone can be used to record handwriting, the microphone to record speech samples, and the camera to record patient movements.

### Other

There are additional devices apart from the ones already mentioned which have also been used to assess a PD patient. We classify devices that do not fall into any of the above categories as “other”. For example, for handwriting assessment, digitized tablets are used as a smart screen to write on during spiralography exams, and smart pens are used to record hand movement during writing. Virtual Reality and Augmented Reality-based solutions have also started becoming popular in PD applications.

## IV. Paper Selection and Classification Methodology

This section presents our methodology to select and classify papers that use technology for PD research. Figure 2 shows an overview of the selection and classification process. We start with an automated search of articles from PubMed Central, Science Direct, IEEE Xplore, and MDPI databases. First, articles that are not relevant to our study are eliminated. For example, articles that include non-human subjects are excluded. In the next step, any duplicate articles that are included in the pool of papers are removed. Finally, the remaining articles are manually classified into one of the four application categories introduced in Section II. In addition, we also mark the mobile technology used in each study according to the categories presented in Section III. We describe each of these steps in more detail in the following sections.

**Fig. 2:**
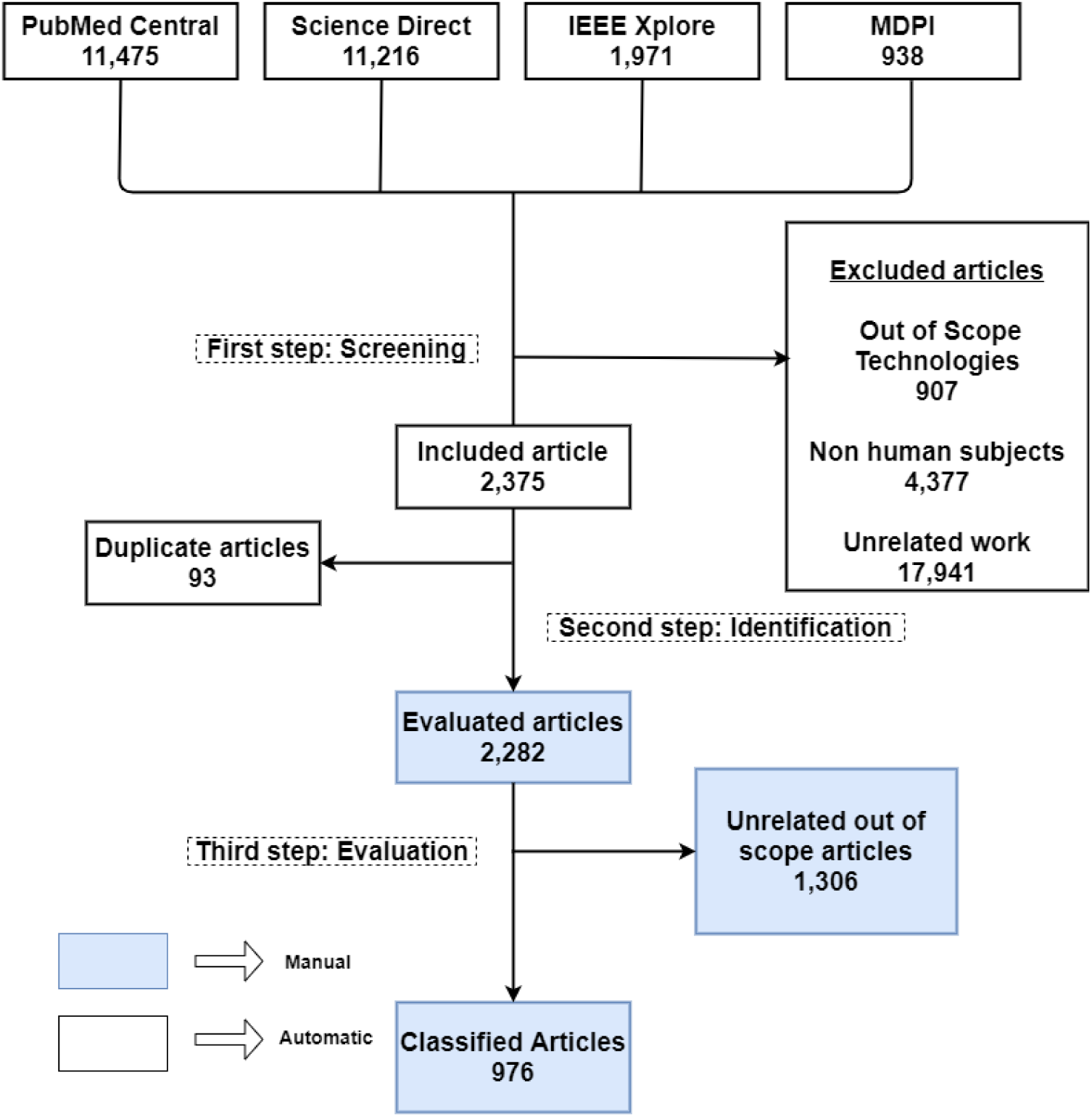
Flow diagram of the systematic review process to new technologies used in assessment of Parkinson’s Disease in the last ten years

### A. Search Methodology

The first step in the review is to obtain articles that studied PD from 2008 to 2019. To this end, we perform an electronic database search of articles published between January 1, 2008, and December 31, 2019, in the PubMed Central, Science Direct, IEEE Xplore, and MDPI databases. These databases are chosen to allow both medical and engineering journals to be included in the search process [36]. The systematic search is performed by following the PRISMA guidelines [22]. The search query includes just the keyword “Parkinson” to keep the search broad. The title/abstract/keyword and year filters are used to provide more specificity. Moreover, only original articles published in English between January 2008 and December 2019, related to Parkinson’s Disease are included in the pool of papers.

The search queries and the number of hits in each database are shown in Table II. A total of 31,940 articles are obtained from the search query, of which 13,070 are from PubMed Central, 16,292 are from Science Direct, 1971 are from IEEE Xplore, and 938 articles are from MDPI. Next, the articles that are not in English or are review articles themselves are removed from the pool of papers. 25,600 articles were left after the process of excluding non-English and review articles. The Title, Authors, Publication Title, Year of Publication (YOP), Keywords, Abstract, Keyword, and DOI are accumulated for all of the 25,600 entries. This database is fed to the filtering step to exclude articles that are out of our scope.

**TABLE II:** Search queries used for each database

| Database | Query | Years | Hits |
| --- | --- | --- | --- |
| Pubmed Central | parkinson[Body - Key Terms] OR parkinson[Abstract]<br>OR parkinson[Title] | 2008 - 2019 | 13070 |
| Science Direct | "parkinson" in Abstract OR Keyword OR Title | 2008 - 2019 | 16292 |
| IEEE Xplore | "parkinson" in Abstract OR Keyword OR Title | 2008 - 2019 | 1971 |
| MDPI | "parkinson" in Abstract OR Keyword OR Title | 2008 - 2019 | 938 |

### B. Filtering Methodology

Information collected from the four databases is used as input for an automated filtering script written in Python. Based on the PICO strategy [22], the script uses four keyword blocks adapted from [37] to implement the selection strategies and the exclusion criteria. The first keyword block has ten keywords related to the disease, followed by the second block of 7 keywords to exclude studies conducted on non-human subjects, as shown in Figure 3. The next set of keyword blocks filter the papers based on the devices used. Specifically, the third keyword block has 66 technology devices commonly used for Parkinson’s Disease assessment. Finally, the last keyword is used to exclude 11 technologies that are unsuitable outside the clinic use. The excluded technologies include MRI, deep brain simulation, PET, neuroimaging, and SPECT. The complete list of all keywords is provided in Appendix C.

**Fig. 3:**
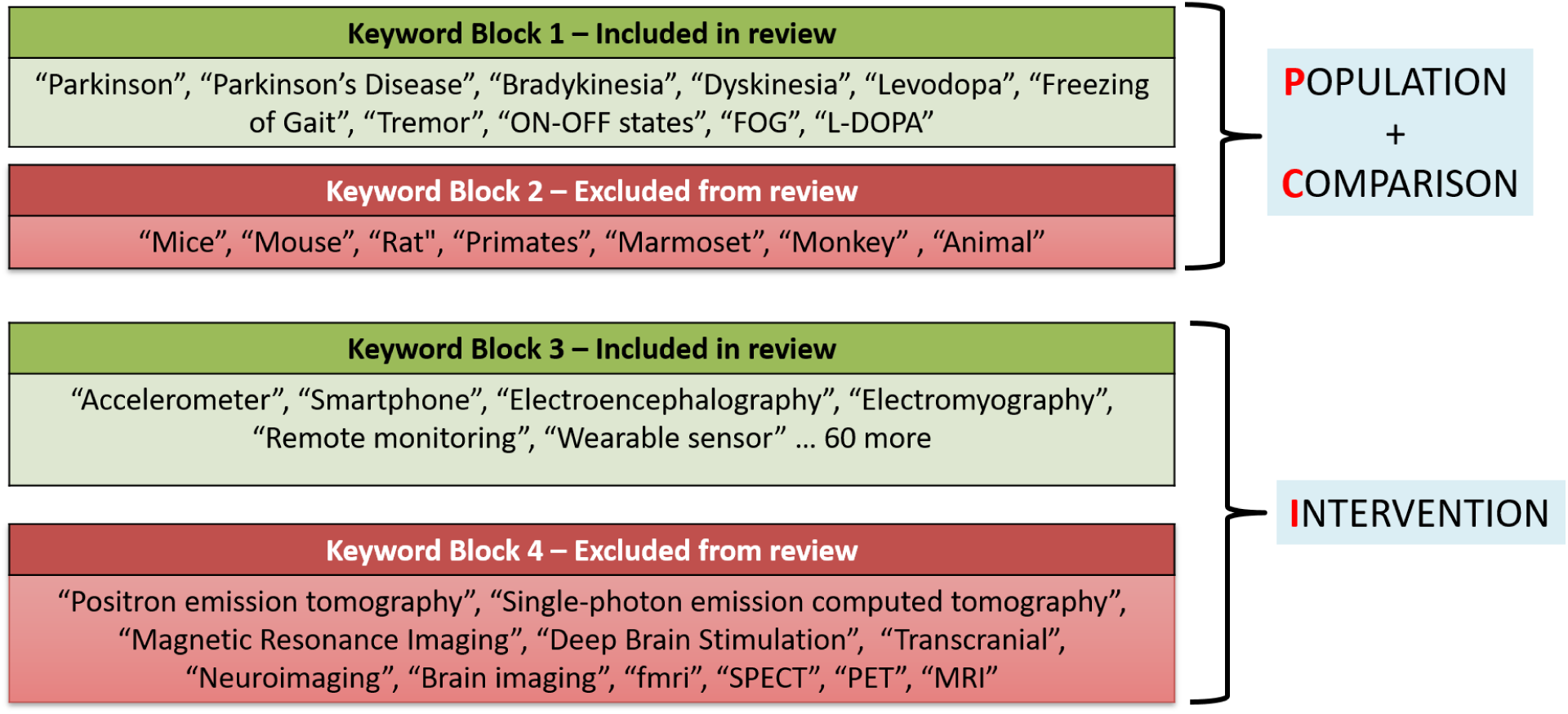
Keyword blocks constructed according to PICOS strategy to determine relevance of a paper to this review

A Python script is used to automate the filtering process. The script goes through each article’s Title, Abstract, and Keywords (TAK). It compares them with the four keyword blocks to objectively determine if the material is relevant to our review or can be excluded, as shown in Figure 3. The first keyword block is applied at the beginning of the process. If the TAK search indicates that the article is not related to PD, further evaluations are skipped. The second keyword block is then applied to exclude studies with non-human subjects, such as rats or monkeys. This keyword block also determines the clinical feasibility of the studies. Similarly, the third and the fourth keyword blocks are used on the remaining list of articles to select or exclude them based on the devices used.

The automated filtering process excluded 23,225 articles from the complete list of 25,600 articles. Specifically, the first keyword block included 7,659 articles relevant to the symptoms that we consider in this review. The second keyword block excluded 4377 articles from the 7,659 articles since they use non-human subjects, thus leaving 3,282 articles for further filtering. Similarly, the third and fourth keyword blocks excluded 907 articles as they involved studies with technologies that are out of our scope. The full list of the keywords are provided in Appendix C. Finally, the script removes duplicate entries, leaving 2282 articles.

The Python code used for filtering and removing the duplicate articles will be made available to the public. Sharing the tools and data used in this study will facilitate future work with more articles published after 2019.

### C. Classification Methodology

The articles obtained after the automated filtering process are manually analyzed to classify the application area and technology used in each study. We first read the title, abstract, and keywords to understand the main application area. Specifically, we choose one of the four application areas described in Section II. We also denote the symptoms (Table I) analyzed by the articles. If the application area or symptoms cannot be extracted from the TAK review, we read the entire paper to classify the application area and the symptoms accurately. The devices and technology used by the studies are also categorized using a similar procedure.

As part of the manual classification, we also exclude any articles that are out of scope. For example, some articles use specific terms, such as sensor and acceleration, although they are out of scope. Similarly, some articles that use “ECG” to monitor “Wolff-Parkinson-White (WPW) syndrome” gets accidentally included by the script, even though Wolff-Parkinson-White (WPW) syndrome is not related to PD. We manually exclude such articles from the final list of classified items. Overall, after manual inspection 976 out of 25,600 articles were identified as relevant and are classified into the application areas and technology used.

## V. Results and Discussion

This section analyzes the research trends between 2008 and 2019 using the classification results from Section IV-C. Before describing the details, we first analyze the overall trend of publications during this period, as shown in Figure 4. The total number of papers published every year has steadily increased since 2008. Figure 4 also shows that the rate of increase has significantly grown in the last four years, i.e., from 2016 to 2019. Notably, most of the papers (62%) were published in the last four years (between 2016 - 2019), while 21% papers in just 2019. This trend shows the growing interest in finding alternative objective ways of assessing Parkinson’s Disease, particularly in the most recent years.

**Fig. 4:**
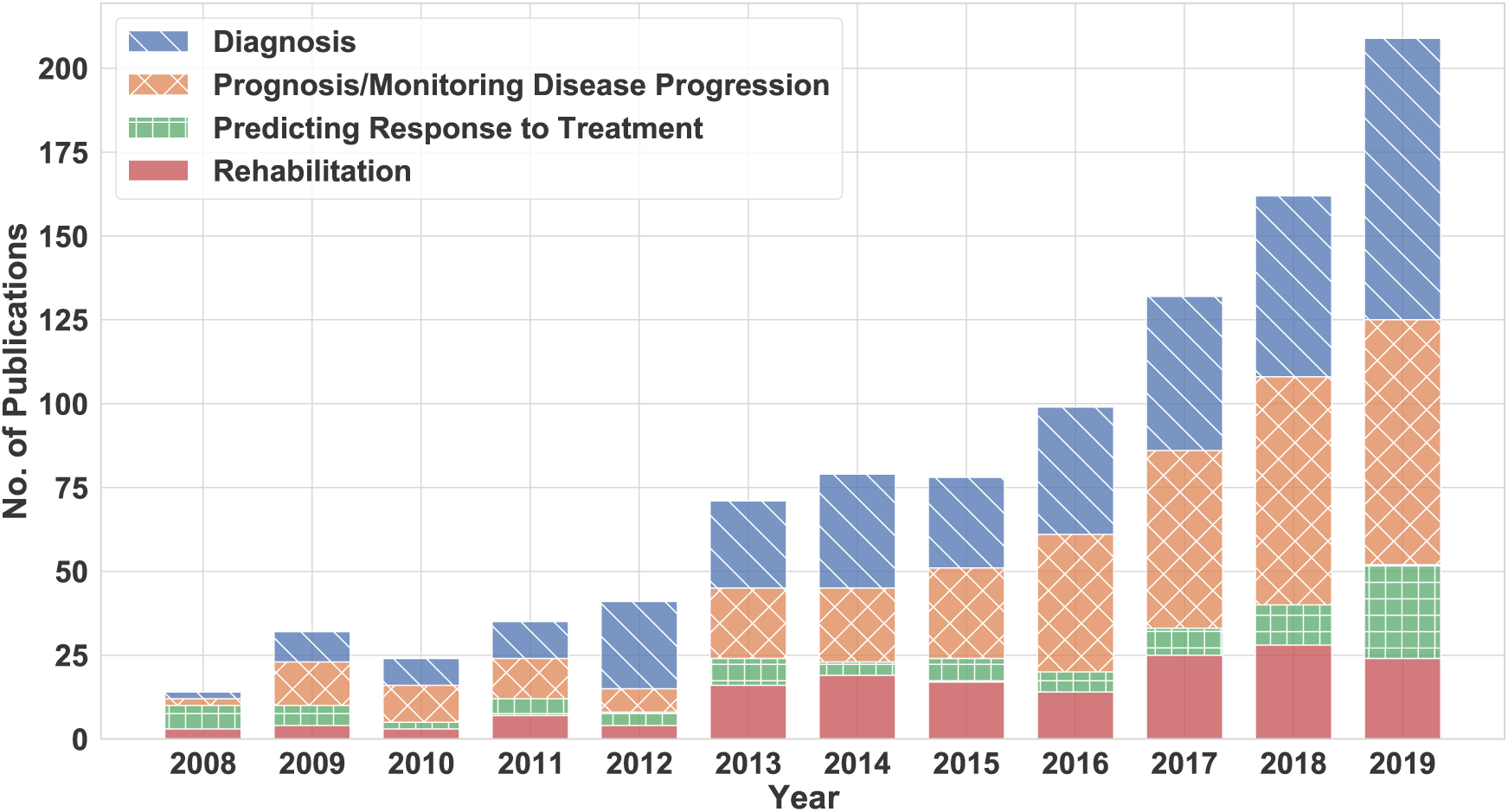
Number of publications using new technologies between 2008-2019

The following subsections first analyze the number of papers published in each application area. Then, we focus on the various PD symptoms studied by these papers. Finally, we describe the various wearable and mobile technologies used for PD diagnosis, monitoring, and treatment. The analysis in this section can help researchers understand the change in PD research over the last few years and future directions in this area.

### A. Application Areas

Wearable technology has varying levels of utility in each application area of PD. To this end, Figure 5 shows the percentage of papers published in each of the four applications used in this survey. Out of the 976 papers evaluated, 365 (37%) studies focus on diagnosis or assisting in the diagnosis of Parkinson’s Disease and 350 (36%) papers are focused on monitoring of patients or prognosis of the disease and the severity of symptoms. Furthermore, 164 (17%) studies are about improving the rehabilitation process of the patients and improving their quality of life, and while 97 (10%) studies analyze the effect of the medication and treatment on the symptoms of the patient as shown in Figure 5. These numbers show that wearable and mobile technology are most frequently used for diagnosis and monitoring of PD. In the following, we detail the trends in each application area.

**Fig. 5:**
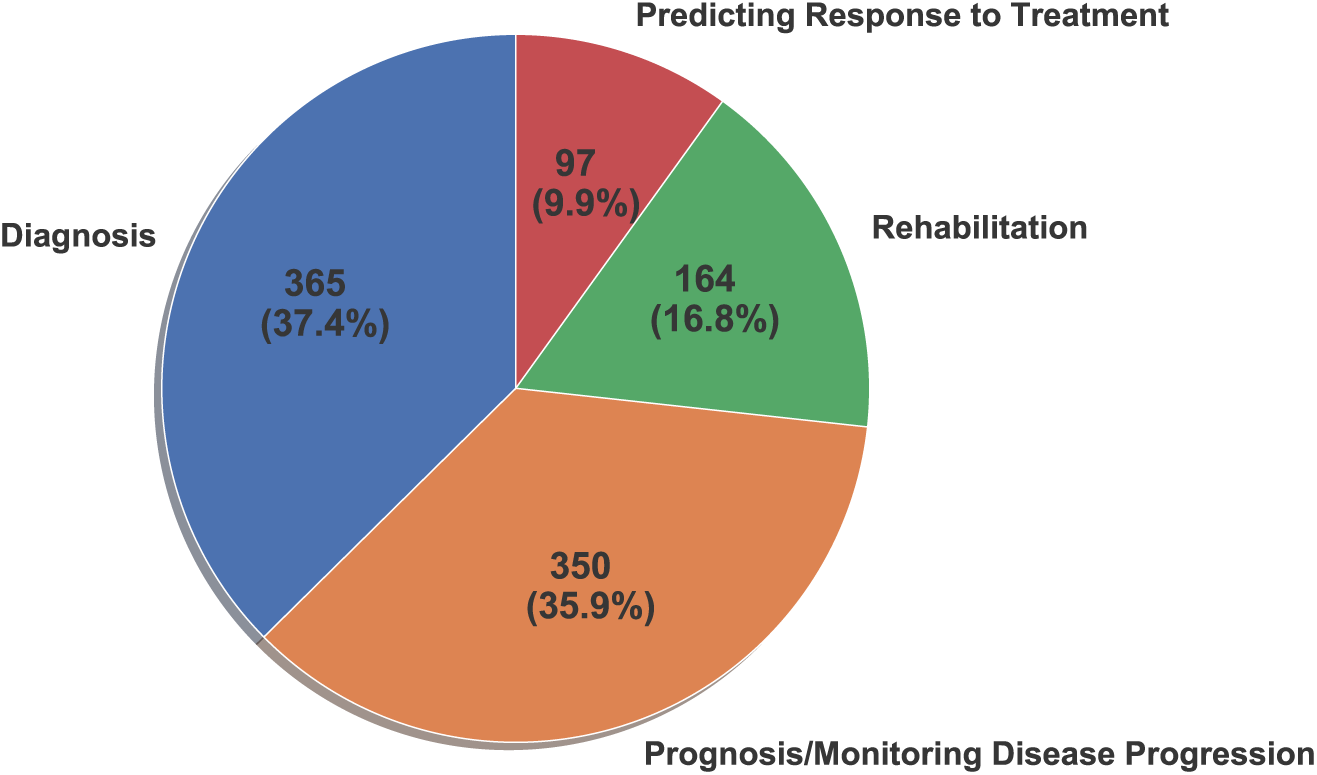
Percentage of publications between 2008-2019 by application area

#### 1) Diagnosis

Since a correct diagnosis is of prime importance, several researchers proposed objective and inexpensive techniques based on mobile technology to augment current techniques. Figure 6(a) shows the trend in the research papers focusing on diagnosis over the last 12 years. We observe a steep increase in the number of articles focusing on the diagnosis of PD. More detailed investigation indicates that studies analyzing both motor and non-motor symptoms contributed to this increase. Raethjen et al. [38] and Zhang et al. [39] have used EEG and EMG data to characterize the tremor in PD patients. Similarly, gait analysis using smartphone accelerometers is used to differentiate PD patients with gait disorder from healthy controls [25]. Muscle activity information obtained from EMG sensors has also been used for PD diagnosis. For example, Meigal et al. [26] employ surface EMG (sEMG) to differentiate between PD subjects from healthy controls and analyze the severity of symptoms. Studies focusing on non-motor symptoms typically analyze sleep patterns and speech disorders as PD biomarkers. EMG data recorded from the chin of a sleeping PD patient to compare the rapid eye movement (REM) sleep chin EMG quantitative features between PD patients with or without REM sleep behavior disorder (RBD) [27]. Similarly, Campos-Roca et al. extracted various acoustic features from an acoustic data set of 40 healthy control and 40 PD patients [28]. In another study, Tsanas et al. used four parsimonious subsets of 132 dysphonia features from an existing data set of 263 samples from 43 subjects [32]. The authors show that the classification with the new dysphonia features was able to reach almost 99% accuracy.

**Fig. 6:**
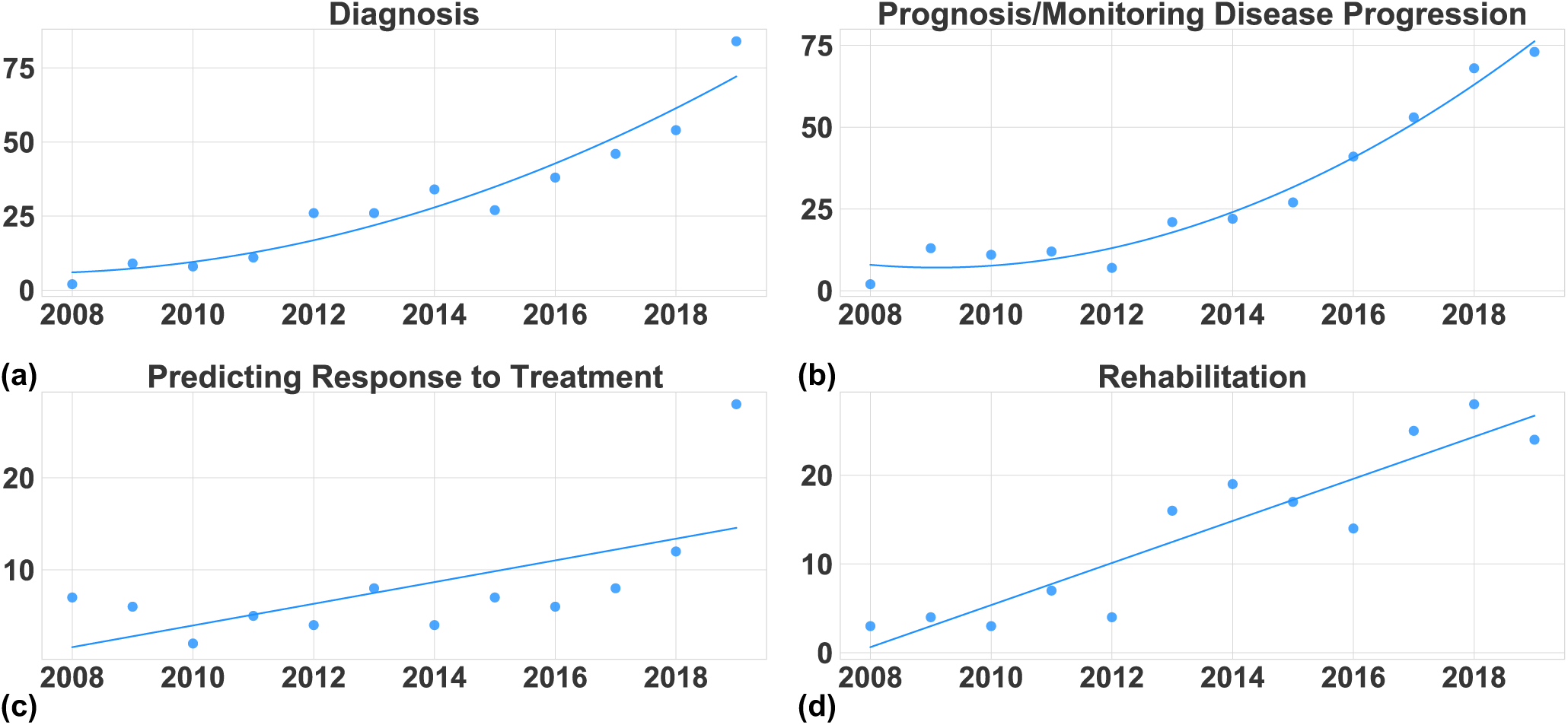
Articles published in last 12 years by PD application area. (a) Publications that are focusing on contributing to improving Diagnosis of PD, (b) Publications that are focusing on Prognosis and Monitoring of PD, its symptoms and assessing severity, (c) Publications that are focusing on Predicting Response to Treatment, (d) Publications that are focusing on improving Therapy and Rehabilitation techniques

In summary, these studies show that recent advances in technology combined with novel algorithms can help clinicians in developing objective measures for PD diagnosis. Table VI in Appendix-B provides a comprehensive list of the articles focusing on diagnosis for interested readers.

#### 2) Prognosis/Monitoring Disease Progression

The assessment of the condition and severity of PD symptoms depends primarily on the clinician’s judgment and the patient’s feedback. However, patient feedback is subject to recall bias and subjectivity, affecting clinician’s analysis. Therefore, there is a strong need for objective home-based monitoring systems to provide feedback to both doctors and patients. Wearable devices and technologies are well suited for home-based monitoring since they can continuously record the patients’ movements and symptoms. 350 (36%) papers published between 2008 and 2019 dealt with problems related to tracking PD patients, assisted living for patients, evaluating the severity of PD symptoms in a patient, and disease progression in general, as shown in Figure 5. Figure 6(b) shows the trend of papers published in the prognosis application area from 2008 to 2019. The number of publications starts increasing considerably faster after 2014. This growth is mainly driven by the improvements in wearable technologies in the last 3-4 years. We provide a brief overview of recent research that uses wearable devices for PD prognosis while Table V in Appendix-B provides a comprehensive list of papers.

The papers in the prognosis application typically focus on monitoring gait parameters. For instance, Zwartjes et al. present an ambulatory monitoring system that provides a complete motor assessment of a PD patient by simultaneously analyzing motor activities and the severity of several symptoms like tremor, bradykinesia, and hypokinesia [35]. Another study in 2016 proposes to quantifying tremors in Parkinson’s patients using smartwatches [40]. Similarly, Bächlin et al. propose a wearable assistant to detect FoG events during ambulatory movements [41]. In addition to monitoring, papers in the prognosis area have also focused on enabling clinical tests outside the clinic. For example, the “Timed Up and Go” test (TUG) is a commonly used clinical test to evaluate balance and mobility. Salarian et al. proposed an instrumented TUG called iTUG using portable inertial sensors [42]. More recently, researchers have focused on non-motor symptoms, such as emotions and fatigue, to assess the progression of PD. Other studies recorded facial expressions of 40 PD patients to investigate the relationship between reduced facial expressiveness and altered emotion recognition in PD [33]. Overall, these studies demonstrate the potential offered by advances in technology to enable monitoring of PD patients. Hence, the trend in Figure 6(b) is likely to continue to grow in the coming years.

#### 3) Predicting Response to Treatment

About 10% of articles classified in Section IV-C measure the efficacy of the PD treatment in a patient. Figure 6(c) shows the trend of the number of papers published each year for predicting the response to therapy. The growth in this application area has been steady over the past decade. In general, this shows that the focus on using wearable and mobile devices for predicting response to PD treatment has not changed significantly from 2008 to 2018. We also observe a steep increase in 2019 in the number of papers focusing on predicting response to treatment. This jump may indicate a growing interest in the research community in predicting response to treatment. Table III shows a summary of the papers that focus on predicting response to treatment. At the same time, the rest of this section highlights the major treatments for PD and technology for predicting the response.

**TABLE III:**
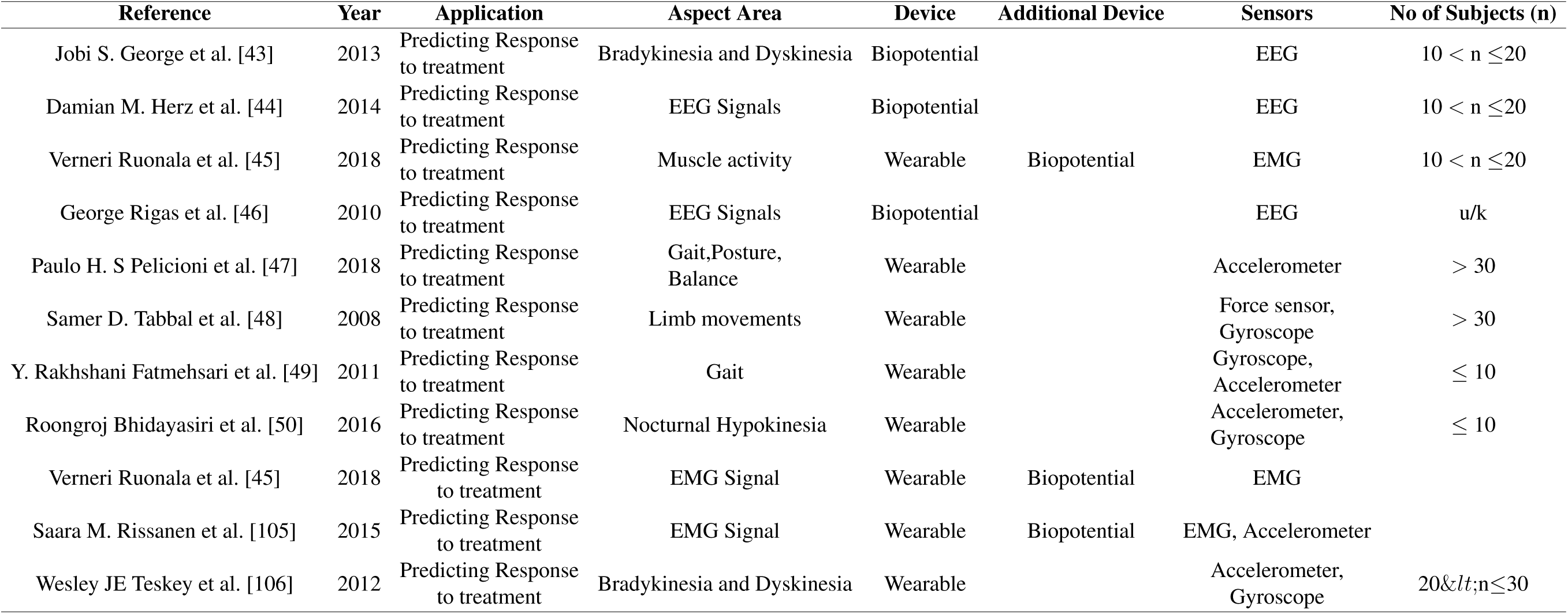
Papers about Predicting Response to Treatment

Levodopa medication is the most popular treatment for motor and non-motor symptoms of Parkinson’s Disease. It helps alleviate important symptoms, such as bradykinesia, rigidity, and tremors, of PD patients. Another conventional treatment methodology used by physicians is Deep Brain Stimulation (DBS) of the subthalamic nucleus (STN). It helps in alleviating motor symptoms and reducing dopaminergic medication. Measuring the efficacy of such treatments on PD patients is important since the mechanism through which they improve cognitive or motor operations in patients with Parkinson’s Disease is not well understood. Recent studies propose theories to measure the effect of Levodopa medication using biopotential devices like EEG and EMG [43]–[45]. In contrast, Rigas et al. [46] and Pelicioni et al. [47] propose new methods of assessing PD symptoms while using wearable sensors. Moreover, other research articles like [48], [49] and [50] have proposed solutions using wearable sensors to measure the effectiveness of DBS treatment on PD patients.

In summary, most of the papers that focus on predicting response to treatment analyze the severity of PD symptoms before, during, and after treatment. Some of the studies also focus on understanding the mechanism through which the medication alleviates PD symptoms. Further research in this domain will help health professionals fine-tune treatment for each patient based on their response to the treatment.

#### 4) Rehabilitation

Developing an efficient rehabilitation plan for a PD patient is crucial to manage the symptoms experienced by patients. Rehabilitation techniques may use assistive cues to help the patients in their daily activities or improve their movement. In the last 12 years, 164 papers have focused on developing therapy and rehabilitation techniques. Significant research attention has also been devoted to developing methods to alleviate motor symptoms like Freezing of gait (FoG) and tremor using auditory, haptic or vibratory cues. Table IV provides a list of papers related to Rehabilitation while we summarize recent approaches in the rest of this subsection.

**TABLE IV:**
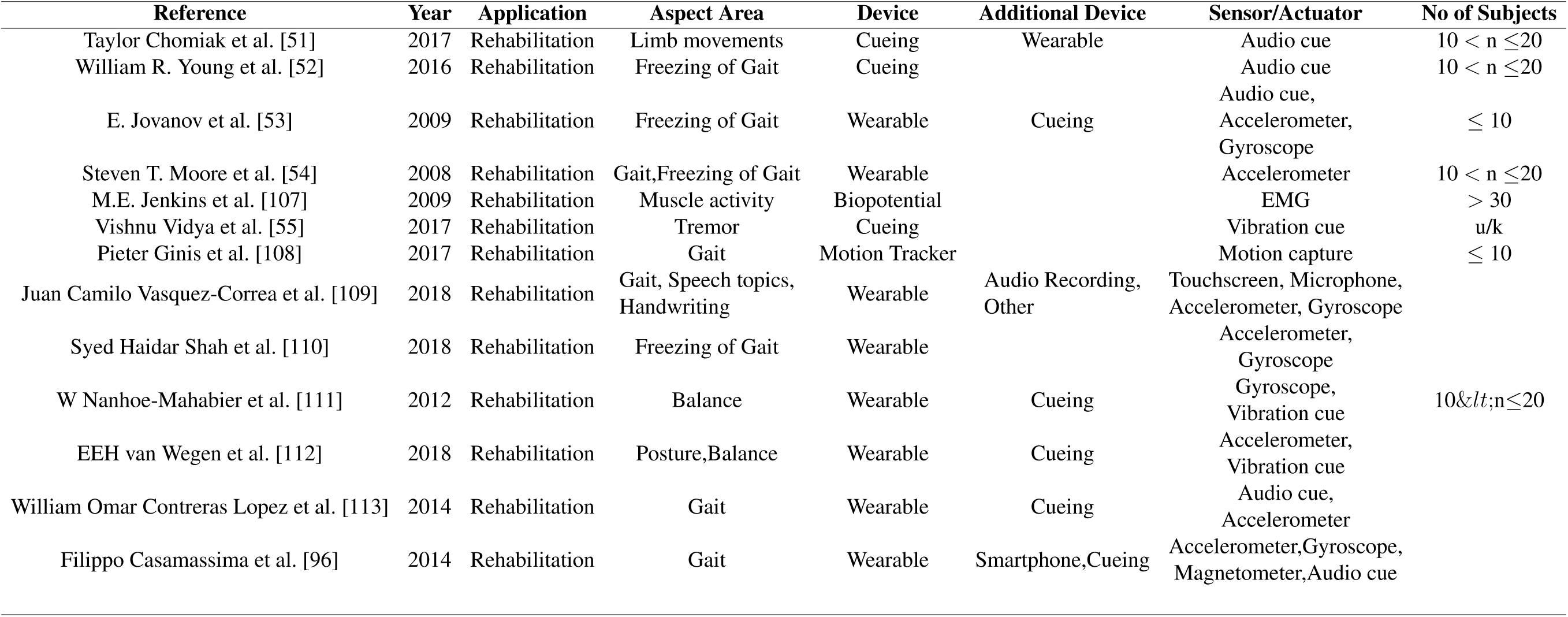
Papers about Rehabilitation

The decline in motor and cognitive functionalities in PD patients leads to increased risk of falling, Freezing of gait, and reduced quality of life. To help with the decline in motor functions, stepping in place (SIP) training is a common rehabilitation technique [51]. Sensory cueing is another common method used for facilitating SIP for PD patients. Recorded sounds of action relevant tasks effectively reduce gait variability in PD patients without Freezing of gait (nFoG). Young et al. studied the efficacy of such auditory cues in PD patients with FoG [52]. Similarly, haptic cues are also used in various studies to unfreeze the gait in FoG episodes for PD patients [53], [54]. To cope with hand tremor, Vidya et al. [55] use a coin-type vibration motor on the patients’ wrist and a micro-controller for generating random vibration patterns by using Pulse Width Modulation.

In summary, the focus of researchers on improving rehabilitation strategies for PD patients has steadily increased since 2008, as shown in Figure 6(d). The growth is linear with the number of papers increasing from about two in 2008 to more than 25 in 2019.

### B. Trends in Symptoms Measured by PD Research Papers

A majority of exploration in the last decade has been in assessing motor symptoms since they are primarily visible in patients. Inertial data collected from IMUs in wearable devices are widely used to monitor gait parameters, tremor, motor activities, FoG events, bradykinesia, and dyskinesia (ON/OFF Stages). Force and pressure sensors placed under the shoe or in an insole sensor are used for measuring the ground reaction force, which is a popular parameter for analyzing gait. EMG sensors are used for monitoring the muscular response of a person. More sophisticated instruments like digitized tablets and smart pens are used to analyze hand movement and pressure while writing. The heat-map in Figure 7 shows the number of times a specific motor symptom is used to develop a solution to a particular application area. Gait abnormality is the most popular motor symptom in PD assessment across all application areas except for “Predicting Response to Treatment”. The motor symptoms that are most commonly monitored among applications focusing on “Predicting Response to Treatment” are bradykinesia and dyskinesia. They primarily monitor the ON/OFF stages and evaluate the muscle activities. Tremor and Freezing of gait are common motor symptoms in a PD patient and are also used as biomarkers for objective assessment of PD. Analyzing the balance and posture of a patient is a common strategy used in prognosis and rehabilitation.

**Fig. 7:**
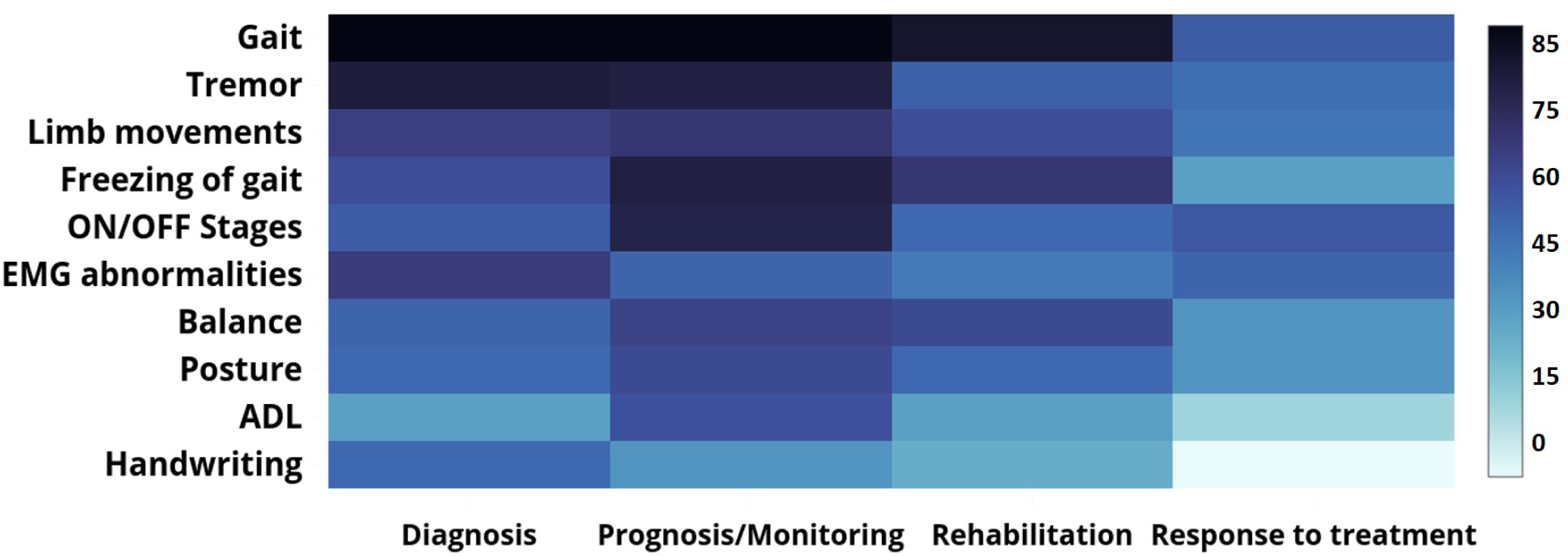
Thermal map indicating the number the publications between 2008-2019 that measure the motor symptoms of PD application areas. The darker the color, the higher the number.

**Fig. 8:**
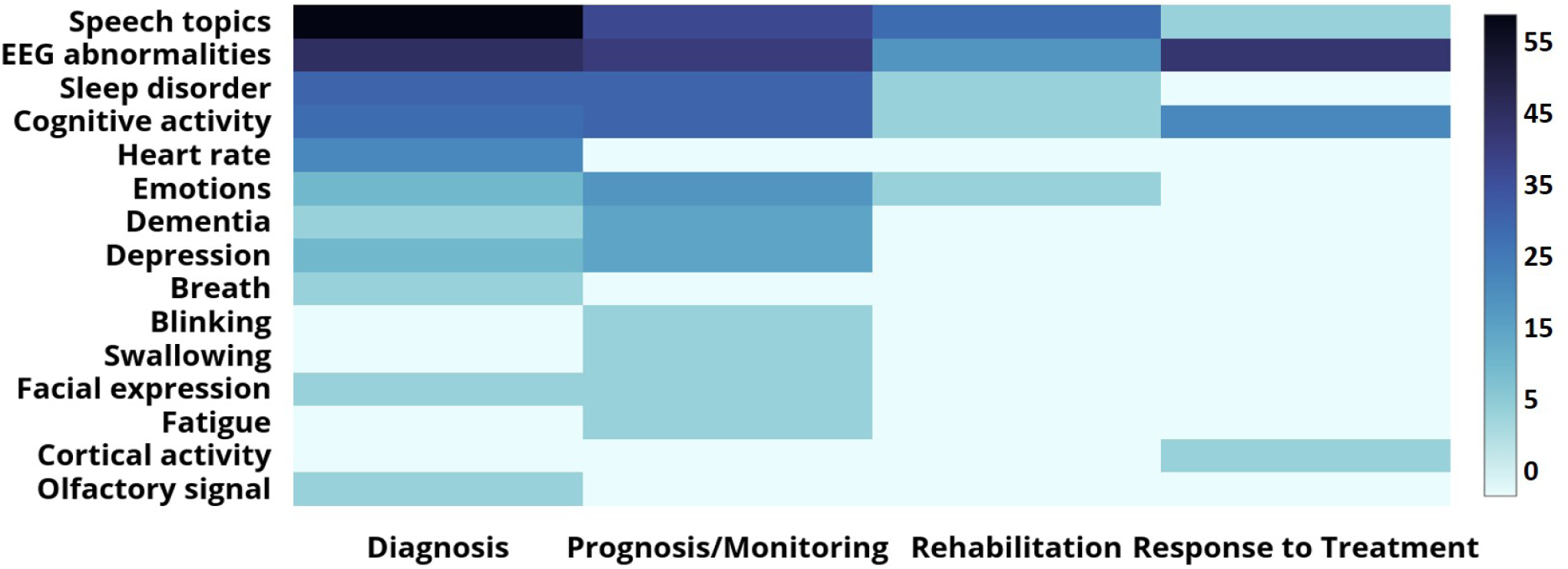
Thermal map indicating the number the publications between 2008-2019 that measure the non-motor symptoms of PD application areas. The darker the color, the higher the number.

Figure 9 shows the trend in which the focus on gait, tremor, and other symptoms has progressed in the last decade. It can be observed that following 2012, more research studies have focused on gait than tremor, even though the number of papers for each of these symptoms kept growing. With the popularity of wearables and with the development of advanced IMUs, measuring of gait parameters became easier.

**Fig. 9:**
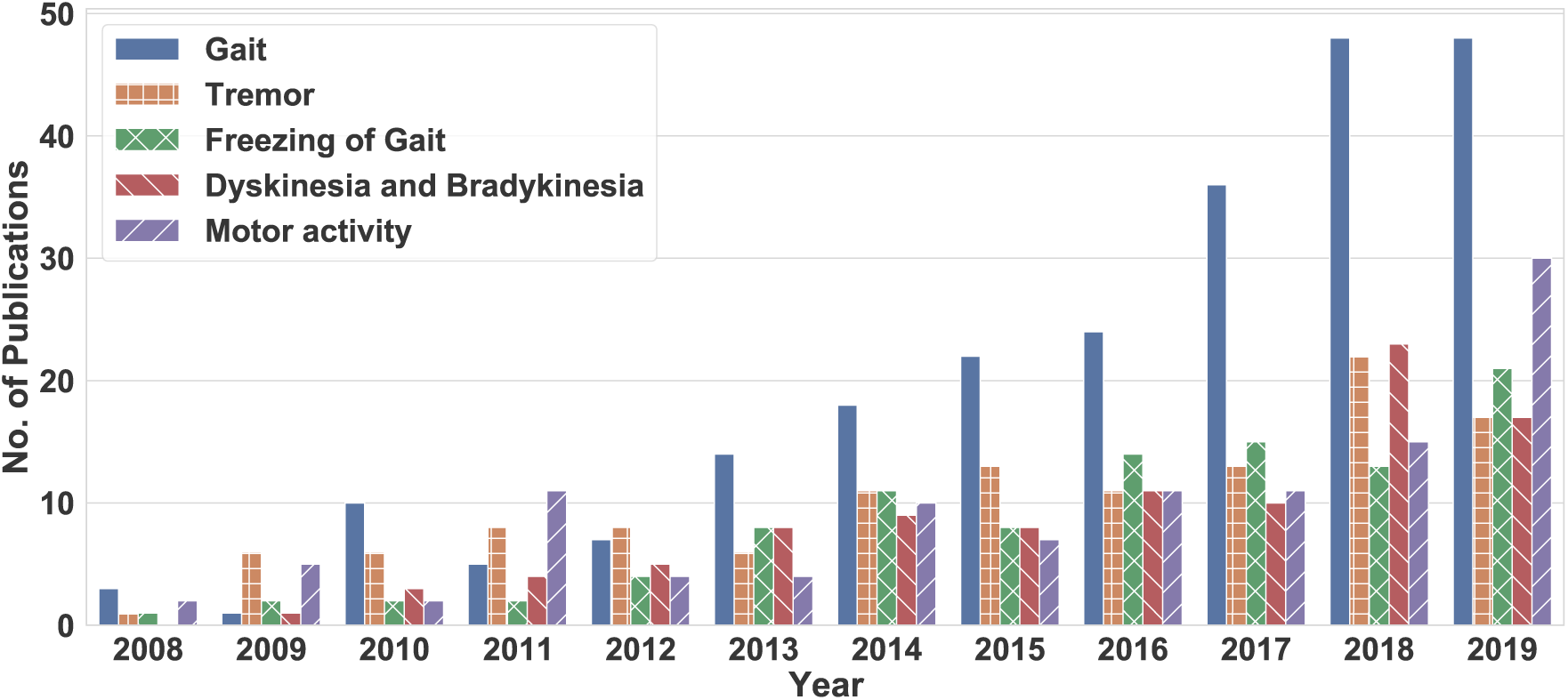
Articles published in last ten years measuring Gait, Tremor, and other symptoms using modern technology

Many researchers are now focusing on non-motor symptoms such as cognitive impairment, dementia, and depression. These can be more disabling for a patient; hence, objective assessment is required. Analyzing neural response measurements is a common strategy used by clinicians and researchers to improve diagnosis, monitoring, and analyzing the response to treatment, as shown in Figure 8. Analyzing the cognitive activity of patients is also a strategy used in different application areas. However, it is evident from the heat-map that the amount of work focusing on severe and disabling symptoms such as dementia, depression, fatigue is negligible compared to the motor symptoms.

### C. Trends in Device Usage from 2008 to 2019

This section evaluates the type of device used in the papers we review. Of the articles reviewed, 44% used a wearable device for collecting the data, 20% papers used biopotential devices, 10% used Audio Recording devices, 65% used Motion Capturing systems, 3% used different Cueing devices like auditory, haptic or visual cues, as shown in Figure 10. The figure shows that the wearable and biopotential devices are the most commonly used devices in the studies related to Parkinson’s assessment.

**Fig. 10:**
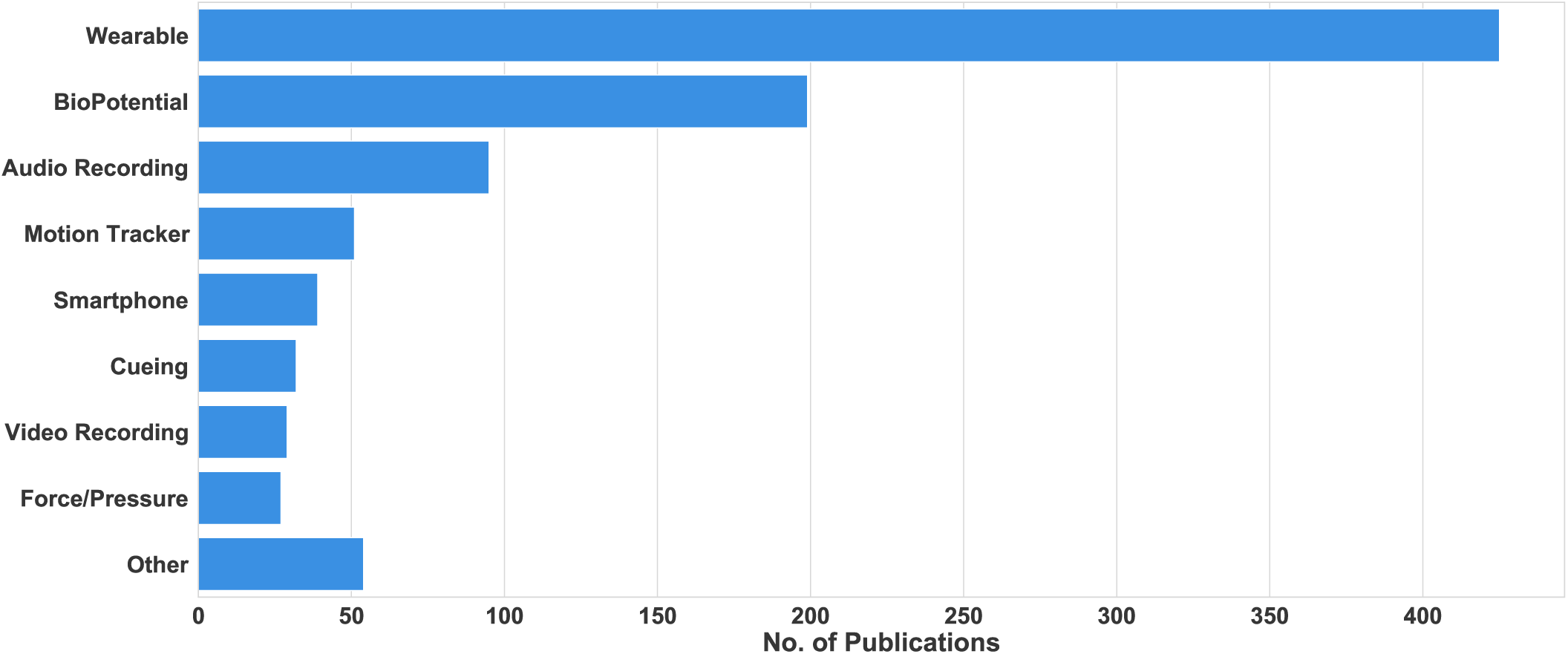
Number of publications using new technologies between the years 2008-2019

Biopotential devices, such as EEG, ECG, EMG, EOG, have been popular in assessing the stage of a patient who has Parkinson’s Disease. EEG recordings are prevalent in measuring the neural activity of a patient, which is a popular PD biomarker [38], [56]–[58]. EEG data is also useful for assessing other non-motor symptoms including sleep topics, dementia, cognitive activity, and mixed symptoms like saccades [59]–[64]. Similarly, EMG recordings are beneficial in analyzing the muscle activity, which is instrumental in detecting and assessing Parkinson’s disease. [65]–[69]. Many researchers also use ECG and EOG to study the heart rate and optical movements, respectively, to assess PD in patients [70]–[72]. The growth in the number of papers using Wearables or Biopotentials is shown in Figure 11(a-b). Figure 11(a) suggests that the number of papers that use biopotential devices to evaluate PD symptoms has grown at a constant rate. With the development of portable biopotential devices and wearable devices with built-in biopotential sensors, their use for PD research is expected to increase.

**Fig. 11:**
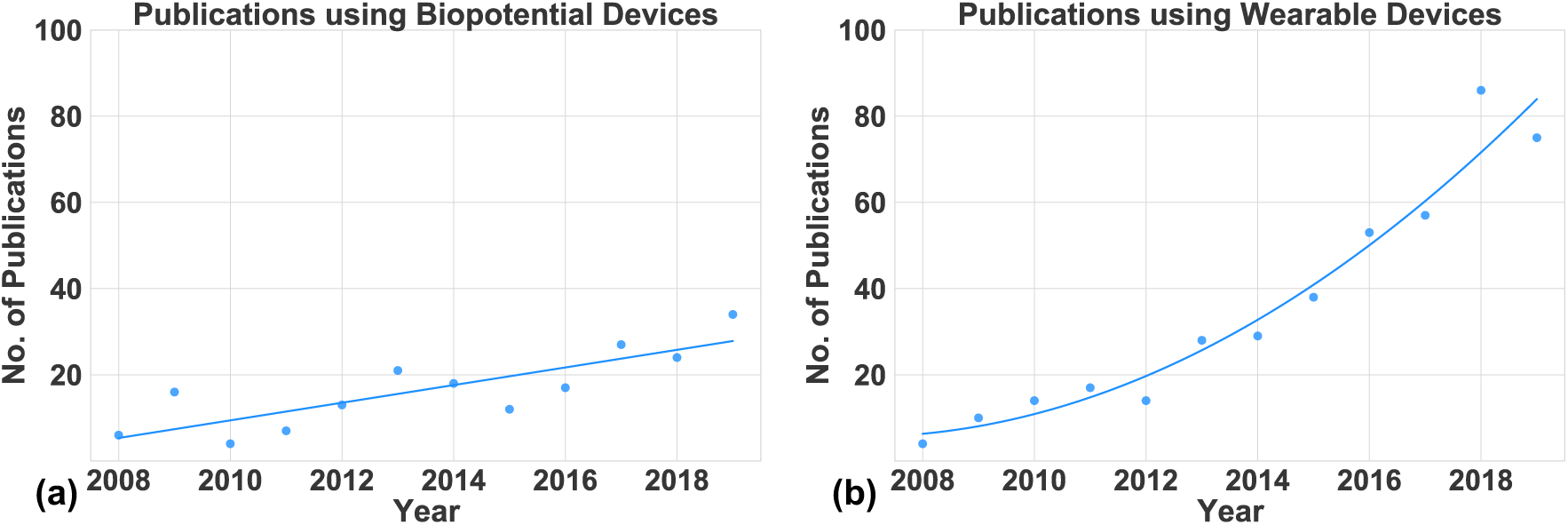
(a) The papers published between 2008 and 2019 that use Biopotential devices for PD assessment. (b) The papers published between 2008 and 2018 that use Wearable devices for PD assessment. The solid line shows the trend of the publications in the last 12 years

Section III elaborated the different devices categorized as “Wearable”. Any wireless device placed on any part of a subject’s body to collect relevant data can be called a wearable device. Such devices usually have one or multiple sensors embedded in them for collecting the data. Then, the raw data is either transmitted via some communication interface, i.e., Bluetooth or Zigbee, to a processing unit or is processed on a microcontroller in the device itself. Different types of sensors, such as accelerometer, gyroscope, magnetometer, temperature, force, and pressure, have been used individually or together for PD assessment [73]–[81]. Wearable devices have also been developed to incorporate Biopotential sensors like EMG used to record muscle activity data [39], [45], [82]–[85]. Other sensors like insole force or pressure sensors have been used to evaluate the vertical ground reaction force generated when the subject is walking to assess their gait, balance, or posture [86]–[92]. Figure 11(b) shows that the number of papers published using wearable devices has increased significantly in the last 11 years and is expected to keep growing.

#### Device Usage Trend

The heatmap in Figure 12 shows the number of papers using each of the devices from 2008–2019. We see that the usage of all the devices has grown in recent years. The increase is significantly pronounced for wearable and biopotential devices. This increase can be attributed to advances in sensor technology, low-power processing, and machine learning algorithms. Motion capturing systems, audio recording, and smartphones are also used more frequently in the last several years, similar to wearable and bipotential devices. They are expected to be used more in the future for home monitoring applications.

**Fig. 12:**
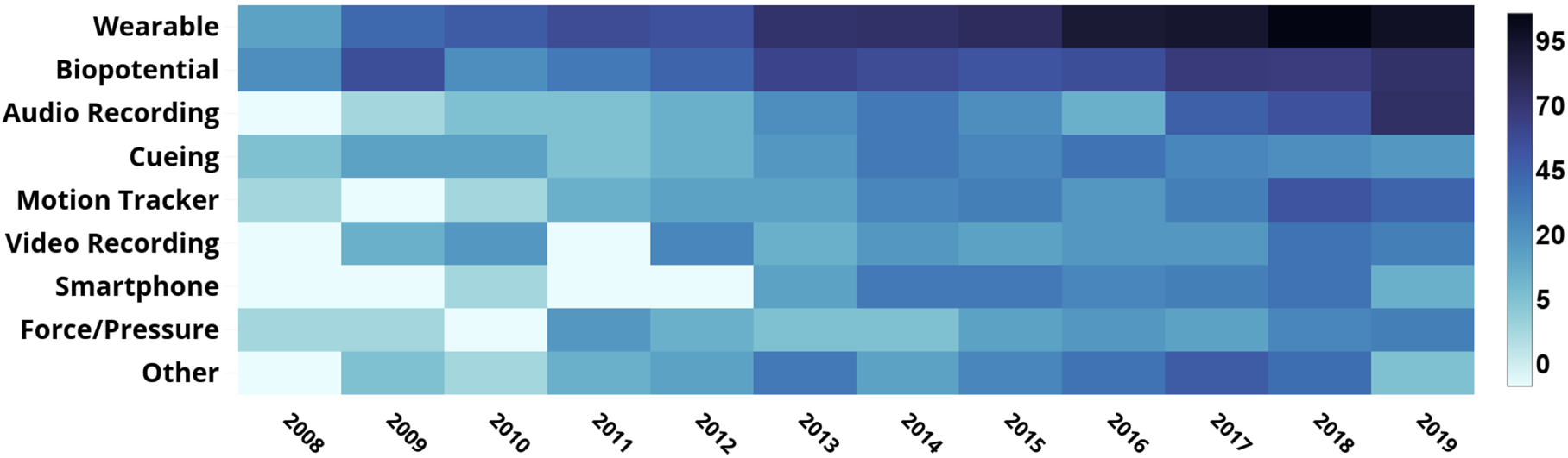
Distribution of articles published between 2008-2019 related to the assessment of Parkinson’s Disease using modern technology

#### Device Usage by Application Area

Figure 13 shows how each device category is used across the four applications. Almost all device categories, except cueing, are predominantly used in diagnosis and prognosis. This trend is expected since a majority of papers in this review focus on diagnosis or prognosis. When the devices are analyzed individually, we see that wearable devices are most commonly used for prognosis due to their ability to monitor patients in a free-living environment. Motion tracking and video recording are also used most commonly for prognosis due to similar reasons. Biopotential devices have one of the highest usages in predicting response to therapy. Audio-recording devices are prevalent in aiding diagnosis but have not been used in predicting response to treatment. Similarly, smartphones are yet to be used in applications focused on predicting response to treatment.

**Fig. 13:**
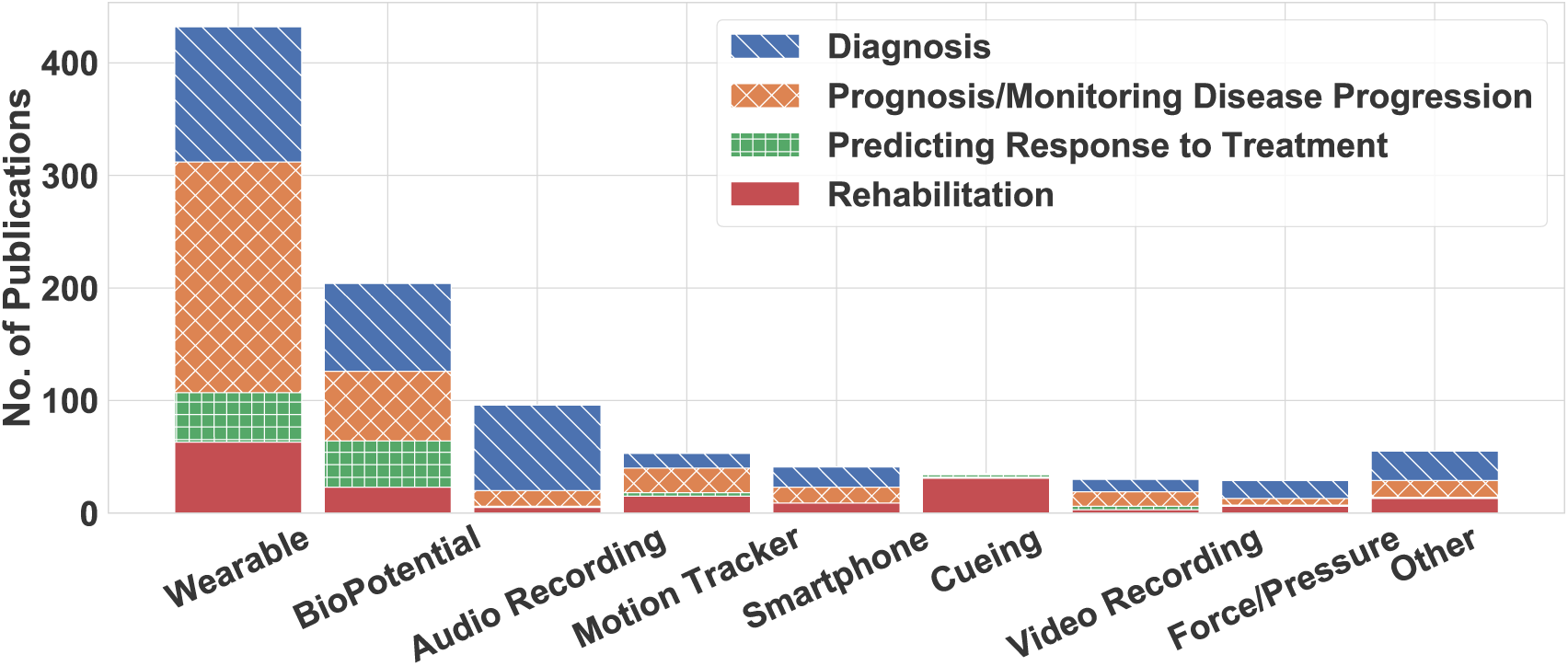
Percentage of publications between 2008-2019 using different novel technologies for PD assessment by application areas

All device modalities have been used for the rehabilitation application. However, cueing devices are the predominant mode for rehabilitation. Specifically, around 90% of the work using cueing systems focuses on improving the therapeutic strategies and rehabilitation plans. Active cueing such as vibration and auditory feedback have also been used to suppress symptoms like FoG and tremor. Different scientific studies have used visual cues to develop strategies for assisting walk of PD patients [93]– [95]. Wireless headphones have been used to give auditory feedback to patients for improved motor activity, gait training, and balance training [94], [96]–[98]. Moreover, auditory and vibratory cues have been used to help a patient break out of a freeze or suppress tremor [41], [53], [55], [99]–[103].

## VI. CONCLUSIONS AND FUTURE WORK

This review presented a comprehensive overview of the technological solutions currently implemented for objective assessment of Parkinson’s Disease and its essential features. We reviewed 976 articles from the last decade to identify four application areas, eight device categories, and the measured symptoms. From this exploratory review, we were able to analyze the trend in which the studies in this field are moving. We conclude that in the scientific community, the emerging idea is to use unobtrusive systems for monitoring the progression of a disease from its nascent stage. One of the limitations of these studies are the limitations of available data sets. The small sample size of subjects reduces the generalizability of the solution and also hampers its credibility. Another limitation is the optimal number and the placement of sensors. Furthermore, there are arguments among the researchers on the correct features that should be extracted from the sensor signals [17]. Lastly, for automatic assessment of the symptoms, efficient algorithms are required to classify the symptoms with high accuracy. Our future work includes developing algorithms for automatic classification of the papers so that we can minimize the need for manual inspection of papers.

## Data Availability

The details of all the evaluated papers are listed in the publicly shared table made available at https://bit.ly/3rcjJgZ

## Author Contributions

Conceptualization, R.D. and U.O.; Methodology, R.D.; Software, R.D., G.B., and S.A.; Validation, R.D., and S.A.; Formal Analysis, R.D.; Investigation, R.D.; Resources, R.D.; Data Curation, R.D. and S.A.; Writing – Original Draft Preparation, R.D., G.B., and S.A.; Writing – Review & Editing, U.O. and H.S.; Visualization, R.D., G.B., and S.A.; Supervision, U.O. and H.S.; Project Administration, U.O.; Funding Acquisition, U.O.

## Acknowledgments

This is a short text to acknowledge the contributions of specific colleagues, institutions, or agencies that aided the efforts of the authors.

## Appendix A

### Data Collection

The methodologies used for downloading the data from the four online databases are also different. The documents were exported in comma-separated values (CSV) format from IEEE Xplore, tab-delimited format from MDPI and BibTex format from Science Direct. While, for PubMed Central, we used a Python-based API, Metapub [104] for an automated search. The information extracted from all of the databases were accumulated and stored together in a .CSV file.

## Appendix B

### List of Papers in Each Application Area

Tables III–VI below lists the paper in each application area discussed in Section V-A.

**TABLE V:** Papers about Prognosis or Monitoring of Disease Progression & Severity of Symptoms

| Reference | Year | Application | Aspect Area | Device | Additional Device | Sensor/Actuator | No of Subjects |
| --- | --- | --- | --- | --- | --- | --- | --- |
| Daphne G. M. Zwartjes et al. [35] | 2010 | Prognosis/Monitoring Disease Progression | Limb movements, Tremor, Movement disorder | Wearable | Video Recording | Accelerometer, Gyroscope, Video camera | $10 < n \leq 20$ |
| R. Contreras et al. [40] | 2016 | Prognosis/Monitoring Disease Progression | Tremor | Wearable | Smartphone | Accelerometer, Gyroscope | $10 < n \leq 20$ |
| Shaohua Wan et al. [114] | 2018 | Prognosis/Monitoring Disease Progression | Speech topics, ADL | Smartphone |  | Accelerometer, Microphone | uk |
| Luis A. Sanchez-Perez et al. [115] | 2018 | Prognosis/Monitoring Disease Progression | Tremor | Wearable | | Accelerometer, Gyroscope, Magnetometer | $> 30$ |
| M Bächlin et al. [41] | 2010 | Prognosis/Monitoring Disease Progression | Gait, Freezing of Gait | Wearable | Cueing | Accelerometer, Audio cue | $\leq 10$ |
| Àngels Bayés et al. [116] | 2018 | Prognosis/Monitoring Disease Progression | Bradykinesia and Dyskinesia | Wearable | Smartphone | Accelerometer | u/k |
| Arash Salarian et al. [108] | 2010 | Prognosis/Monitoring Disease Progression | Posture, Gait | Wearable | | Accelerometer, Gyroscope | $10 < n \leq 20$ |
| JA Robichaud et al. [117] | 2009 | Prognosis/Monitoring Disease Progression | EMG Signal | Biopotential | | EMG | $20 < n \leq 30$ |
| Mevludin Memedi et al. [118] | 2015 | Prognosis/Monitoring Disease Progression | Handwriting | Other | | Touchscreen | $> 30$ |
| J Dietz et al. [119] | 2013 | Prognosis/Monitoring Disease Progression | Emotions, Depression | Biopotential | | EEG | $10 < n \leq 20$ |
| Lucia Ricciardi et al. [33] | 2015 | Prognosis/Monitoring Disease Progression | Emotions | Video Recording | | Video camera | $> 30$ |
| Shyamal Patel et al. [34] | 2009 | Prognosis/Monitoring Disease Progression | Tremor, Bradykinesia and Dyskinesia | Wearable | Video Recording | Accelerometer, Video camera | $10 < n \leq 20$ |

**TABLE VI:**
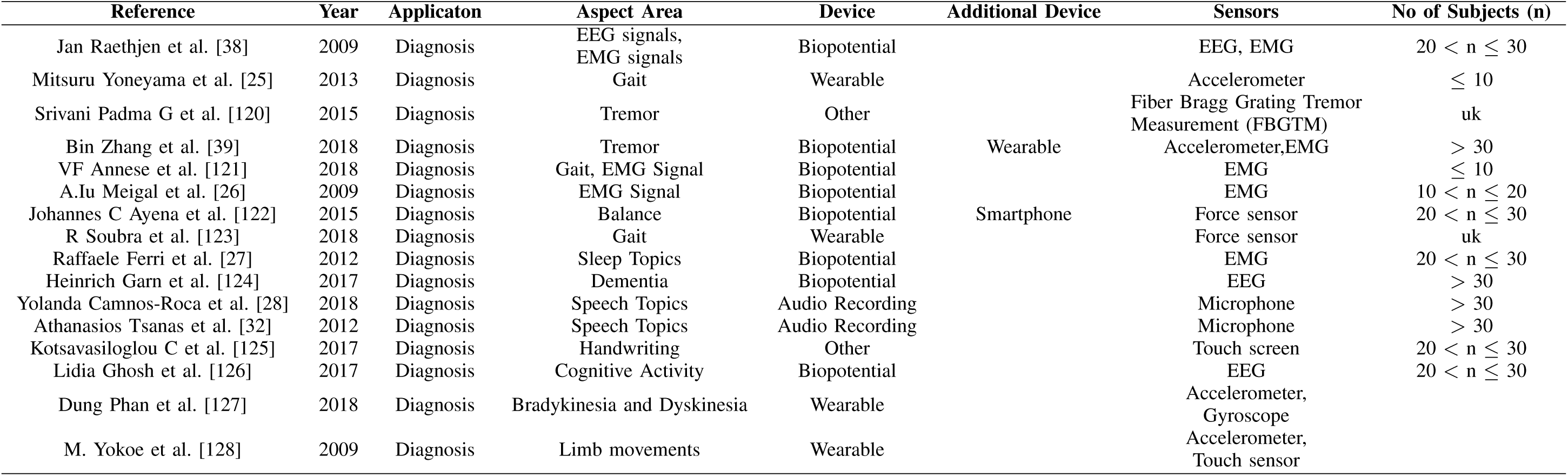
Papers about Diagnosis, Early Diagnosis and Differential Diagnosis

## Appendix C

### Keywords used for Automated Filtering

**The first keyword block has 10 keywords related to the disease:** *“Parkinson”, “Parkinson’s Disease”, “Bradykinesia”, “Dyskinesia”, “Levodopa”, “Freezing of gait”*.

**The second block is made of 7 keywords to exclude non-human studies from the study:** *“Rats”, “Primates”, “Marmoset”, “Monkeys”, “Mice”, “Mouse”, “Animal”*.

**The third keyword block has 66 technology terms related to Parkinson’s Disease assessment:** (i.e., *“Acceleration”, “Accelerometer”, “Gyroscope”, “Magnetometer”, “Gyro”, “Acc”, “Exoskeleton device”, “Inertial sensor”, “Video recording”, “Video camera”, “Camera”, “EHealth”, “Technology”, “Remote monitoring”, “Home monitoring”, “Telemedicine”, “Mobile phone”, “Mobile application”, “Precision medicine”, “Digital health”, “Inertial Measurement Unit”, “IMU”, “Wearable”, “Magneto inertial sensor”, “Sensor”, “Force plate”, “Force sensor”, “Pressure sensor”, “Gait mat”, “Smartphone”, “Augmented reality”, “Kinect”, “VICON”, “ECG”, “EEG”, “EMG”, “EOG”, “MEG”, “Electrocardiography”, “Electroencephalography”, “Electromyography”, “Electrooculography”, “Magnetoencephalography”, “Machine learning”, “Classification”, “Auditory cue”, “Auditory cueing”, “Visual cue”, “Ha[tic cue”, “Vibration”, “Gait classification”, “Insole sensors”, “Leap motion”, “Computer vision”, “Supervised learning”, “Neural networks”, “Speech disorder”, “Music therapy”, “Smart application”, “Smartphone application”, “Smart watch”, “Smart pen”, “Touchscreen”, “Digitized tablet”, “Fitness band”, “Optical motion capturing system”*).

**The last keyword block has 11 technology terms related to Parkinson’s Disease treatment, which we have excluded from our review** (i.e., *“Positron emission tomography”, “Single-photon emission computed tomography”, “Magnetic Resonance Imaging”, “Deep Brain Stimulation”, “Transcranial”, “Neuroimaging”, “Brain imaging”, “fmri”, “SPECT”, “PET”, “MRI”*). The keyword blocks were used based on an earlier review [37]

